# Social inequalities in pregnancy metabolic profile: findings from the multi-ethnic Born in Bradford cohort study

**DOI:** 10.1101/2024.02.08.24302335

**Authors:** Ahmed Elhakeem, Gemma L Clayton, Ana G Soares, Kurt Taylor, Léa Maitre, Gillian Santorelli, John Wright, Deborah A Lawlor, Martine Vrijheid

## Abstract

**Background:** Lower socioeconomic position (SEP) is associated with adverse pregnancy and perinatal outcomes and with less favourable metabolic profile in nonpregnant adults. However, socioeconomic differences in pregnancy metabolic profile are unknown. We investigated association between a composite measure of SEP and pregnancy metabolic profile in White European (WE) and South Asian (SA) women.

**Methods:** We included 3,905 WE and 4,404 SA pregnant women from a population-based UK cohort. Latent class analysis was applied to nineteen individual, household, and area-based SEP indicators (collected by questionnaires or linkage to residential address) to derive a composite SEP latent variable. Targeted nuclear magnetic resonance spectroscopy was used to determine 148 metabolic traits from mid-pregnancy serum samples. Associations between SEP and metabolic traits were examined using linear regressions adjusted for gestational age and weighted by latent class probabilities. An interactive application was developed for exploring all association results (https://aelhak.shinyapps.io/SEP_NMR_BiB/).

**Results:** Five SEP sub-groups were identified and labelled ‘Highest SEP’ (48% WE and 52% SA), ‘High-Medium SEP’ (77% and 23%), ‘Medium SEP’ (56% and 44%) ‘Low-Medium SEP’ (21% and 79%), and ‘Lowest SEP’ (52% and 48%). Lower SEP was associated with more adverse levels of 113 metabolic traits, including lower high-density lipoprotein (HDL) and higher triglycerides and very low-density lipoprotein (VLDL) traits. For example, mean standardized difference (95%CI) in *concentration of small VLDL particles* (vs. Highest SEP) was 0.12 standard deviation (SD) units (0.05 to 0.20) for ‘Medium SEP’ and 0.25*SD* (0.18 to 0.32) for ‘Lowest SEP’. There was statistical evidence of ethnic differences in associations of SEP with 31 traits, primarily characterised by stronger associations in WE women e.g., mean difference in *HDL cholesterol* in WE and SA women respectively (vs. Highest-SEP) was - 0.30*SD* (−0.41 to −0.20) and −0.16*SD* (−0.27 to −0.05) for ‘Medium SEP’, and −0.62*SD* (−0.72 to −0.52) and −0.29*SD* (−0.40 to −0.20) for ‘Lowest SEP’.

**Conclusions:** We found widespread socioeconomic differences in metabolic traits in pregnant WE and SA women residing in the UK, and clearer socioeconomic gradient for some traits in WE women. Supporting all pregnant women in the most disadvantaged socioeconomic groups may provide the greatest benefit for perinatal health.

## BACKGROUND

Extensive changes in maternal circulating metabolites occur during pregnancy, which are likely to be important for maternal health and normal fetal development^1–3^, with some of these metabolites associating with adverse pregnancy and perinatal outcomes^4–6^. Studies indicate that lower socioeconomic position (SEP) associates with adverse pregnancy and perinatal outcomes, including gestational diabetes, preterm birth, and small-for-gestational-age^7–9^. Lower SEP (indicated by a lower educational level and occupational class) has also been associated with worse metabolic profile in adolescents and adults^10^ however, to the best of our knowledge, SEP differences in pregnancy metabolic profile have not been examined.

Besides SEP, ethnicity differences in pregnancy and perinatal outcomes^9,11–13^ and metabolic traits have been reported^14^. For example, evidence from the Born in Bradford (BiB) cohort shows that South Asian pregnant women had higher levels of amino acids, fatty acids, and glucose, and lower levels of cholesterol and lipoproteins than White Europeans^14^. Findings from BiB also show differences in SEP between White European and South Asian women^15^, and studies report differences between ethnic groups in associations of SEP with pregnancy and perinatal outcomes^7,8^. Understanding socioeconomic differences in pregnancy metabolic profiles, including across ethnic groups, may help inform public health interventions. Further, studies often relate only one or a small number of indicators of SEP to an outcome, and so rarely acknowledge that SEP is multidimensional and reflects different but related factors including education, occupation, income, wealth, assets, and area deprivation^16,17^.

The aim of this study was to examine the associations between a composite measure of SEP, that should better reflect its multidimensional nature, and mid-pregnancy metabolic profiles in White European and South Asian women.

## METHODS

This study was done according to a pre-specified and publicly available analysis plan^18^ and is reported in line with the STROBE guidelines.

### Cohort description

BiB is a population-based prospective pregnancy cohort that included 12,453 women who experienced 13,776 pregnancies between 2007 and 2011^19^. Most women were recruited at approximately 26–28 weeks gestation at their oral glucose tolerance test, which is offered to all women booked for delivery at Bradford Royal Infirmary. BiB has almost an equal split of White European and South Asian women, all residing in Bradford, UK, a city in the North of England with high levels of socioeconomic deprivation (the BiB study was started due to a high prevalence of poor child health in the city). Mothers, and their partners, recruited into the study provided detailed interview questionnaire data, measurements, and biological samples. The study website gives further information, including protocols, information on data access, and a list of all data (https://borninbradford.nhs.uk/research/documents-data/).

For this study, we included all first enrolled pregnancies to White European and South Asian women (the two main ethnic groups in BiB). After excluding other ethnicities, and those with missing data on SEP, metabolic traits, and gestational age at measurement of metabolic traits, our analysis sample comprised of 3,905 White Europeans and 4,404 South Asians (**Figure 1**).

**Figure 1.**
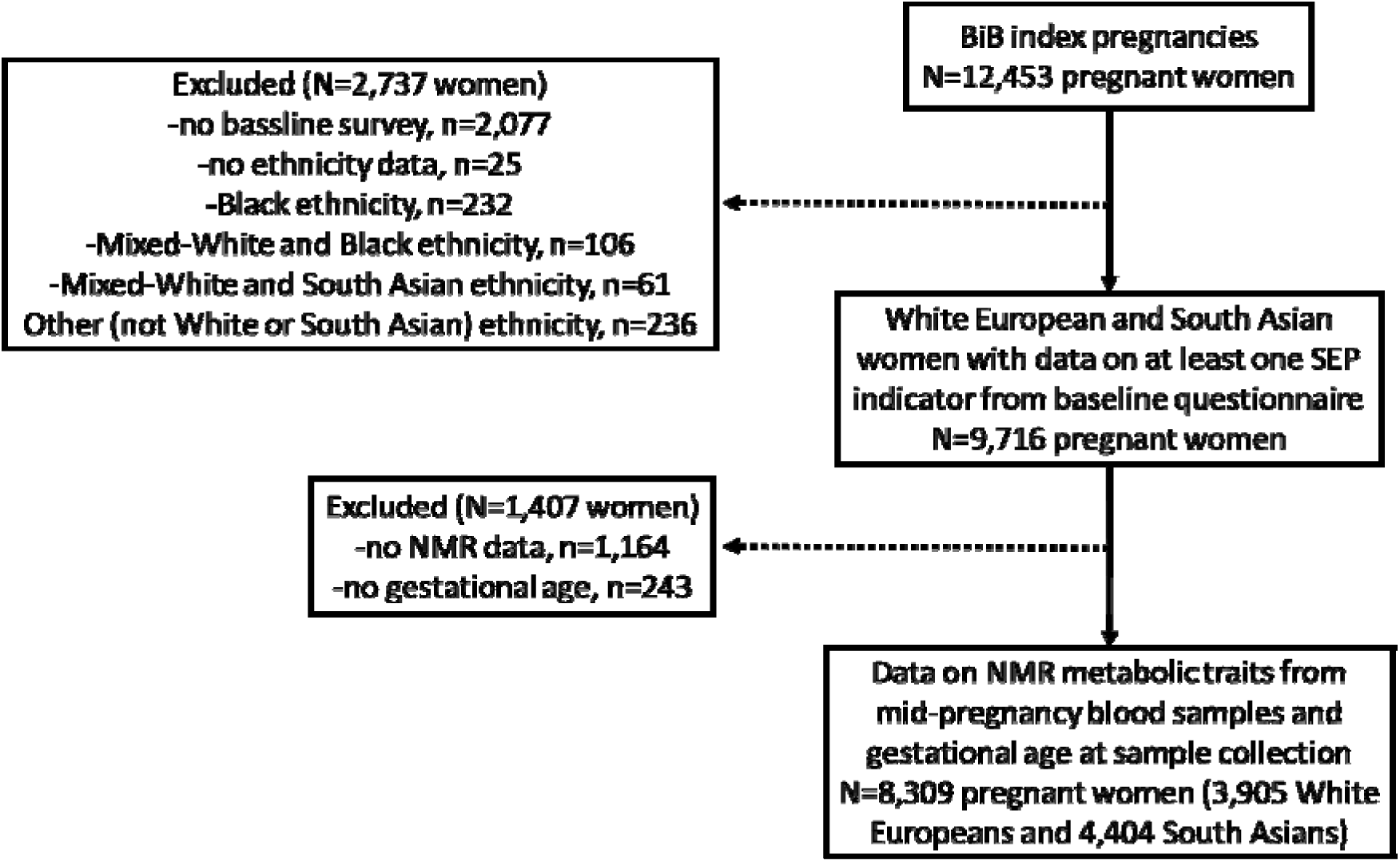
Study flowchart.

### Ethnicity assessment and groups

Ethnicity was reported by the mother at the recruitment questionnaire interview or abstracted from medical records (for those missing questionnaire data) and defined according to the UK Office for National Statistics guidelines. For our main analysis, ethnicity groups were defined as White European or South Asian. White European ethnicity included women that indicated they were White British (n=4,489) or other White European (n=306). South Asian ethnicity included women that indicated they were Pakistani (n=5,128), Indian (n=439), Bangladeshi (n=263) or other South Asian heritage (n=63).

### Indicators of SEP

A total of 19 individual-, household- and area-based indicators of SEP were used to derive a composite SEP latent variable (**Table 1**). All individual- and household-based indicators were reported by the mother using questionnaires in pregnancy, and the area-based indicator was based on linkage to residential address.

**Table 1.** SEP indicators used to derive the composite SEP latent class sub-groups.

| Socioeconomic indicator | White European<br>(n = 3,905) | South Asian<br>(n = 4,404) |
| --- | --- | --- |
| Index of Multiple Deprivation [No. (%)] |  |  |
| Q1 (least deprived) | 1947 (49.9) | 960 (21.8) |
| Q2 | 1118 (28.6) | 1638 (37.2) |
| Q3 (most deprived) | 839 (21.5) | 1805 (41.0) |
| Woman's education [No. (%)] |  |  |
| <5 GCSE equivalent | 733 (21.0) | 1057 (25.3) |
| 5 GCSE equivalent | 1308 (37.5) | 1296 (31.0) |
| A-level equivalent | 675 (19.3) | 568 (13.6) |
| Higher than A-level | 776 (22.2) | 1254 (30.0) |
| Baby's father's education [No. (%)] |  |  |
| <5 GCSE equivalent | 663 (24.1) | 650 (18.9) |
| 5 GCSE equivalent | 1000 (36.4) | 1016 (29.5) |
| A-level equivalent | 470 (17.1) | 418 (12.1) |
| Higher than A-level | 615 (22.4) | 1365 (39.6) |
| Woman's employment status [No. (%)] |  |  |
| Currently employed | 2572 (65.9) | 1211 (27.6) |
| Previously employed | 989 (25.3) | 1269 (28.9) |
| Never employed | 342 (8.8) | 1915 (43.6) |
| Baby's father's employment status [No. (%)] |  |  |
| Non-manual | 1824 (50.8) | 1433 (34.5) |
| Manual | 1068 (29.7) | 1622 (39.0) |
| Self-employed | 368 (10.2) | 825 (19.8) |
| Unemployed | 332 (9.2) | 279 (6.7) |
| Means tested benefit [No. (%)] |  |  |
| Yes | 1383 (35.5) | 1911 (43.5) |
| No | 2508 (64.5) | 2478 (56.5) |
| How well mother and partner managing financially<br>[No. (%)] |  |  |
| Living comfortably | 1036 (26.7) | 1213 (27.7) |
| Doing alright | 1607 (41.3) | 1847 (42.2) |
| Just about getting by | 973 (25.0) | 981 (22.4) |
| Quite difficult or very difficult | 272 (7.0) | 333 (7.6) |
| Financial circumstance compared to year ago [No.<br>(%)] |  |  |
| Better off | 1101 (28.3) | 1308 (30.1) |
| Worse off | 1007 (25.9) | 717 (16.5) |
| About the same | 1778 (45.8) | 2315 (53.3) |
| Able to afford two pairs of all-weather shoes [No. (%)] |  |  |
| Yes | 3611 (96.1) | 4269 (99.3) |
| No | 145 (3.9) | 33 (0.8) |
| Able to afford a small amount of money to spend on yourself each week [No. (%)] |  |  |
| Yes | 2966 (79.8) | 3614 (86.5) |
| No | 751 (20.2) | 565 (13.5) |
| Able to afford to make regular savings of £10 a month [No. (%)] |  |  |
| Yes | 2697 (74.3) | 3294 (80.8) |
| No | 932 (25.7) | 783 (19.2) |
| Housing tenure |  |  |
| Owns outright | 152 (4.2) | 1068 (27.9) |
| Mortgage | 1845 (50.7) | 2020 (52.7) |
| Private landlord | 1050 (28.9) | 484 (12.6) |
| Social housing | 590 (16.3) | 261 (6.8) |
| Overcrowding (person per room) ([No. (%)]) |  |  |
| Q1 | 2548 (65.4) | 1547 (35.2) |
| Q2 | 849 (21.8) | 1393 (31.7) |
| Q3 | 500 (12.8) | 1454 (33.1) |
| Up to date with bills [No. (%)] |  |  |
| Yes | 3369 (88.1) | 3854 (91.4) |
| No | 454 (11.9) | 363 (8.6) |
| Able to afford to replace or repair major electrical goods [No. (%)] |  |  |
| Yes | 2485 (70.2) | 3096 (79.2) |
| No | 1057 (29.8) | 813 (20.8) |
| Able to keep home warm enough in winter [No. (%)] |  |  |
| Yes | 3761 (97.8) | 4175 (96.0) |
| No | 84 (2.2) | 172 (4.0) |
| Able to afford to replace any worn out furniture [No. (%)] |  |  |
| Yes | 2437 (69.1) | 2830 (72.8) |
| No | 1088 (30.9) | 1059 (27.2) |
| Able to afford household contents insurance [No. (%)] |  |  |
| Yes | 2407 (84.6) | 2184 (82.2) |
| No | 438 (15.4) | 474 (17.8) |
| Able to afford to keep home in decent state of decoration [No. (%)] |  |  |
| Yes | 3581 (94.3) | 3743 (88.3) |
| No | 215 (5.7) | 497 (11.7) |

Numbers shown for those with data on metabolic traits and gestational age at blood sample collection). IMD and overcrowding groups were generated in the combined sample.

Area SEP was based on the English Index of Multiple Deprivation (IMD) Score in 2007 (i.e., at the time of pregnancy). IMD is a relative composite measure of multiple deprivation at small area level across England (mean population size in each area is 1500 residents). The domains used to derive IMD in 2007 were income deprivation; employment deprivation; health deprivation and disability; education deprivation; crime deprivation; barriers to housing and services deprivation; and living environment deprivation.

Individual-based indicators reflect educational attainment, occupation, need for financial benefits, and financial circumstances. The highest educational qualification obtained by the woman and the baby’s father was recorded along with the country it was obtained in. We equivalised the highest educational qualifications (based on qualification received and the country obtained) into one of seven categories using the UK National Academic Recognition Information Center. Those with equivalised education coded as other, foreign unknown, or do not know were excluded from each education variable. Because over 25% of women reported that they had never been employed, women’s employment was coded as currently employed, previously employed, or never employed. The baby’s father’s occupation was coded based on the National Statistics Socio-Economic Classification. Those coded as student, unemployed, or don’t know were excluded from this variable.

Women were coded as being in receipt of means tested benefits if they reported receiving any of income support, income tested jobs seekers allowance, working families tax credit, or housing benefit. Women were asked how well they are managing financially with responses being either living comfortably, doing alright, just about getting by, or quite difficult or very difficult. Women were also asked how they are doing financially compared to a year ago, with responses coded as better off, worse off, about the same, or does not wish to answer. Those coded as does not wish to answer were excluded from this variable. Women also reported if they were able to have two pairs of all-weather shoes, money to make regular savings of £10 a month, and a small amount of money to spend each week on themselves.

Household-based indicators reflected questions about housing tenure, overcrowding, and ownership of material items and goods based on questions from the Households Below Average Income Survey. Housing tenure was reported as one of seven groups; owns outright, owns with a mortgage, lives rent free, owned by a private landlord, living in social housing, other and don’t know. Those coded as living rent free, other, or don’t know were excluded from this variable. Responses to questions on numbers of household members and bedrooms were used to derive an indicator of overcrowding based on the person per room approach^20^ by dividing the number of persons by the number of bedrooms in this household. Women were also asked whether they were up to date with household bills, if they had contents insurance, enough money to keep the home in a decent state of repair, money to replace any worn out furniture, money to replace or repair major electrical goods, and if they were able to keep their home warm enough in winter. For each of these variables, women that responded as don’t want/need, doesn’t wish to answer, or don’t know were excluded.

### Pregnancy metabolic traits

Full details of all metabolomic measurements undertaken in BiB have been published^21^. In this study we focus on maternal pregnancy nuclear magnetic resonance (NMR) metabolic traits measured in mid-pregnancy. Women had a fasting mid-pregnancy serum sample taken by trained phlebotomists working in the antenatal clinic of Bradford Royal Infirmary (92% were obtained between 26–28 weeks gestation). Samples were processed within 2.5 hours and placed in −80° freezers. There were no sample freeze-thaw events prior to their use for metabolomic profiling. In total, 227 metabolic traits were measured using a high-throughput targeted NMR platform (Nightingale Health©, Helsinki, Finland). The metabolic traits were quantified in absolute concentration units or ratios and included circulating lipoprotein lipids and subclasses, fatty acids, fatty acid compositions, amino acids, traits related to glycolysis, ketone bodies, fluid balance, and an inflammatory marker^22,23^. In this study, derived measures and ratios were excluded, leaving 148 metabolic traits for analysis (**Additional File 1: Data Set 1**). Gestational age at serum sample collection was recorded.

### Statistical analysis

Latent class analysis (LCA) was applied to all 19 SEP indicators to derive a composite SEP latent variable consisting of SEP sub-groups (latent classes). LCA is a finite mixture model that classifies individuals into unobserved sub-groups (called latent classes) based on their responses to two or more indicator variables, with the aim of identifying subgroups where individuals are more similar within groups than between groups^24^. LCA was done in White European and South Asian women (combined) with at least one SEP indicator and data on pregnancy metabolic traits, and gestational age at metabolic traits’ sample collection. To avoid local maxima solutions, we used 1500 random sets of starting values for the initial stage, 150 final stage optimizations, and 15 initial stage iterations. Models with two to six latent classes were compared and the optimal number of classes was identified based on a combination of BIC, entropy statistic, and Lo-Mendell-Rubin adjusted likelihood ratio test (**Additional File 2: Table S1**). Where these indicators disagreed, the more interpretable model was selected.

Linear regression models with robust standard errors were then used to examine associations between SEP latent class sub-groups (versus a reference SEP sub-group) and each metabolic trait. Models were weighted by the sum of latent class probabilities to allow for uncertainty in SEP latent class membership assignments and were adjusted for gestational age to control for gestational age-related differences in metabolic traits. A SEP by ethnicity (i.e., White European, or South Asian) interaction term was included in all models to investigate ethnic differences in associations between SEP and metabolic traits. All metabolic traits were standardised (by ethnicity group to mean=0, SD=1) to aid comparison of results between different metabolic traits^25^. In sensitivity analysis, we repeated the LCA and regression modelling separately in White European and South Asian women and separately in the White British and Pakistani women (the two biggest groups within the White European and South Asian ethnic groups).

To avoid overloading the main paper, here we present results for selected groups of metabolic trait and sub-particles regardless of *P*-values, and provide all results with exact *P*-values and false discovery rate (FDR) corrected P-values^26^ in additional files. All the results can also be viewed on the accompanying interactive app (https://aelhak.shinyapps.io/SEP_NMR_BiB/). LCA was done in Mplus version 6, and all other analyses were done in R version 4.2.2.

### Missing data

LCA handled missing data on SEP indicators using full information maximum likelihood estimation, with all women with ≥1 SEP indicator variable included in the LCA, under the missing at random assumption (i.e., that the probability of a missing SEP indicator can be entirely explained by other observed SEP indicators and so is not related its value). For the regression analysis, women with missing data on metabolic traits and gestational age were excluded. To explore the potential impact of missing data, we compared characteristics of included women with those excluded due to missing data (**Additional File 3: Table S2**).

### Deviations from pre-specified analysis plan

Following feedback on previous versions of this work presented at scientific conferences and scientific meetings, we decided to make the combined LCA analysis our focus instead of the ethnicity-specific analyses. We decided to analyse traits in SD units instead of perfuming log transformation because virtually all pregnancy metabolic traits were normally distributed (**Additional File 4: Figure S1**). No other changes were made to the analysis plan^18^.

## RESULTS

### Participant characteristics

A total of 3,905 White European and 4,404 South Asian pregnant women with at least one SEP indicator and data on metabolic traits and gestational age at collection of serum samples for the assessment of metabolic traits were included in the study (**Figure 1**). Mean gestational age was 26.6 weeks (SD=1.8) in White Europeans and 26.7 weeks (SD=1.9) in South Asians, and mean age was 26.6 (SD=6.0) and 27.9 (SD=5.2) years respectively. When compared with included women, those excluded due to missing data on metabolic traits and gestational age (n=1,407) had higher proportion of South Asian ethnicity (63% versus 53%) and broadly similar socioeconomic circumstances as indicated by similar levels across most SEP indicators (**Additional File 3: Table S2**).

### SEP sub-groups

LCA (in the combined sample of White Europeans and South Asians) identified five SEP sub-groups which we have labelled ‘*Highest SEP’*, ‘*High-Medium SEP’*, ‘*Medium SEP’*, ‘*Low-Medium SEP’*, and ‘*Lowest SEP’*. The proportions of White Europeans and South Asians in the *Lowest SEP*, *Medium SEP*, and the *Highest SEP* groups were broadly similar, but there were fewer White Europeans than South Asians in the *Low-Medium SEP* group (29% vs. 79%) and more in the *High-Medium SEP* group (77% vs. 23%) (**Figure 2**). The differentiation into SEP sub-groups was driven largely by five SEP indicators (mother’s educational level and employment status, partner’s educational level and occupational class, and means tested benefits). There was little difference between SEP sub-groups in whether women reported being able to keep the home warm enough in winter and being able to afford to afford two pairs of all-weather shoes. The remaining 12 indicators each contributed with modest differences. When compared with *High-Medium SEP* subgroup, the *Low-Medium SEP* sub-group was more likely to own a house outright without a mortgage (**Figure 2**).

**Figure 2.**
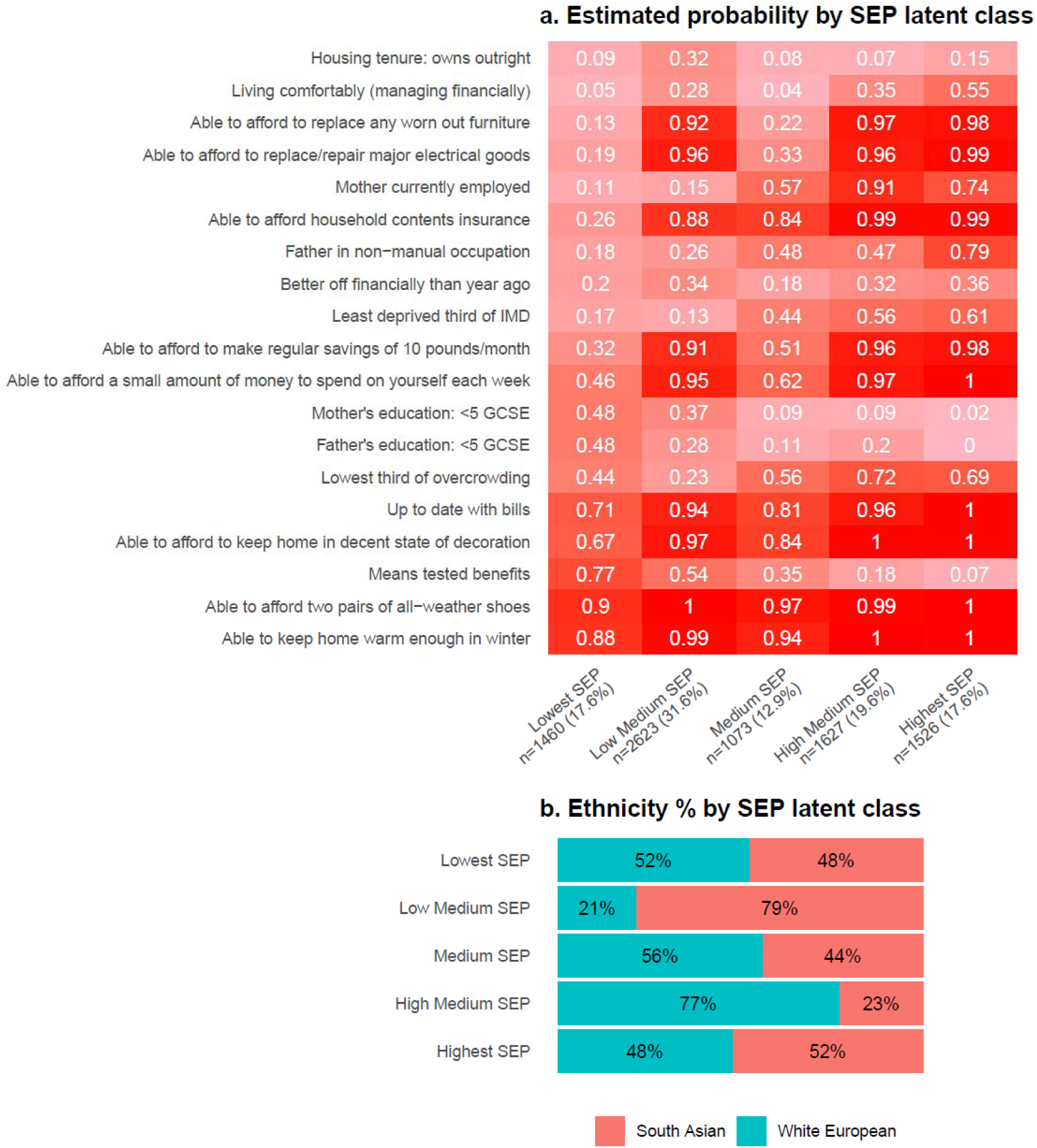
Estimated mean probabilities and the proportions of White European and South Asian women in each SEP sub-group from the combined SEP latent class analysis.

### SEP sub-groups and pregnancy metabolic traits

Lower SEP was associated with (mostly) less favourable levels of 113 metabolic traits at the FDR corrected *P*<0.05 threshold (**Additional File 5: Data Set 2**, **Additional File 6: Data Set 3**). This included associations between lower SEP and higher VLDL cholesterol, total triglycerides (**Figure 3**), glycoprotein acetyls, and VLDL concentration, lower levels of cholines (**Figure 4**), and higher VLDL and lower HDL in cholesterol and phospholipids (**Figure 5**). Conversely, there was less evidence of associations with LDL particles and no differences in albumin, glycine, histidine, or lactate (**Additional File 6: Data Set 3**).

**Figure 3.**
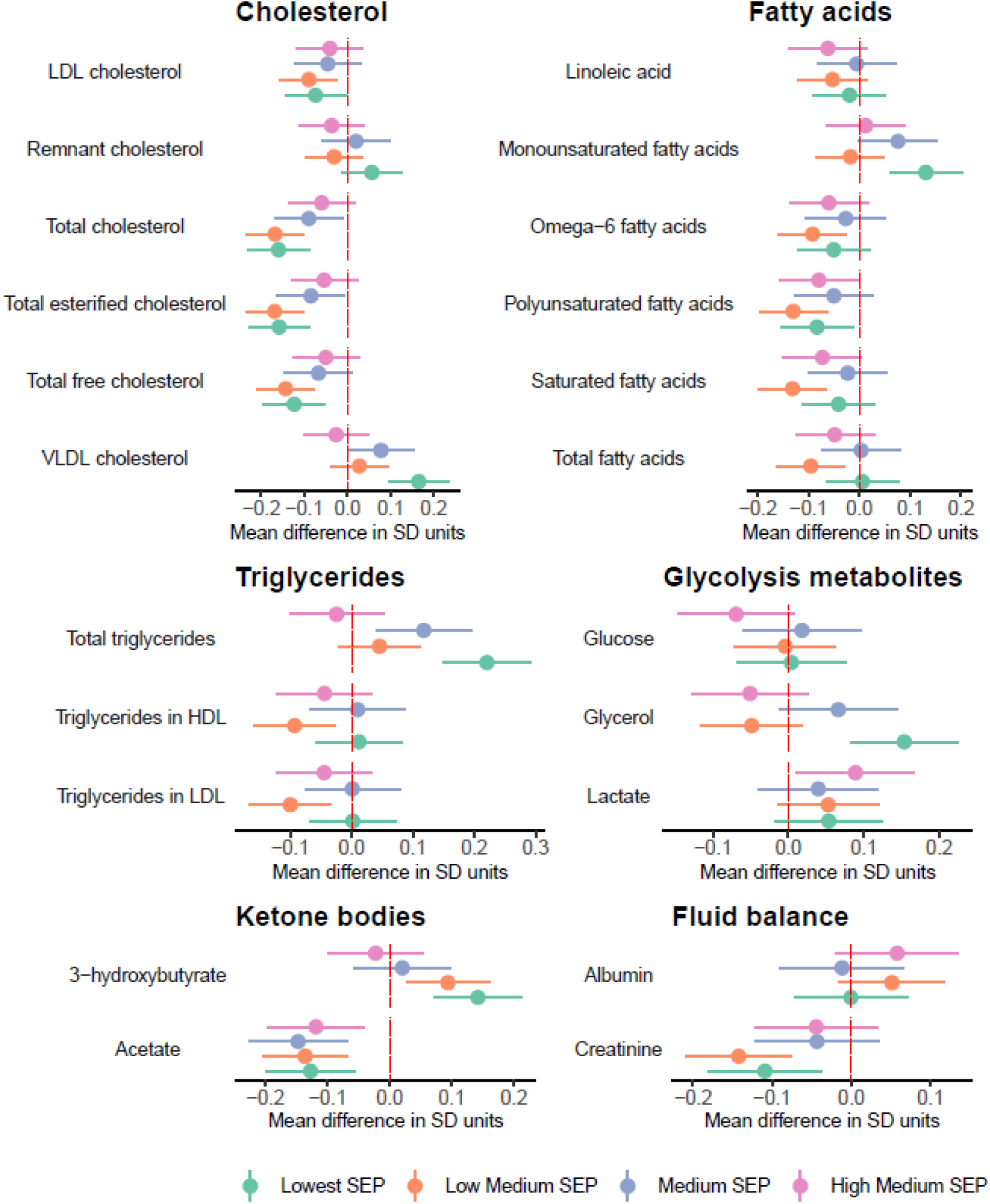
Mean difference in cholesterol, fatty acids, triglycerides, glycolysis-related metabolites, ketone bodies, and fluid balance traits by SEP sub-groups in the combined sample of White European and South Asian women, shown for traits without statistical evidence of SEP by ethnicity interaction (reference: Highest SEP sub-group).

**Figure 4.**
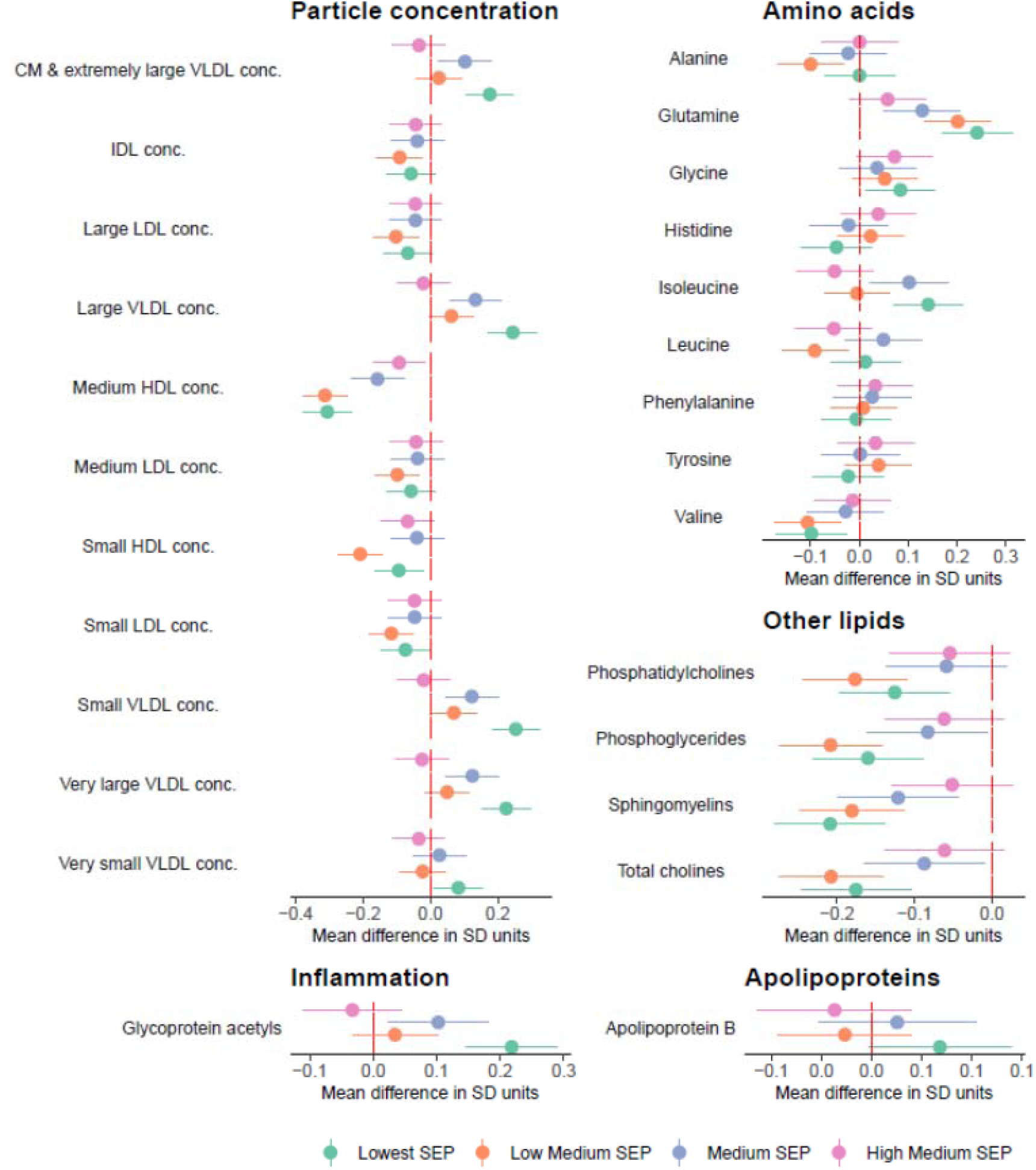
Mean difference in lipoprotein particle concentration, amino acids, other lipids, inflammation, and apolipoproteins by SEP sub-groups in the combined sample of White European and South Asian women, shown for traits without statistical evidence of SEP by ethnicity interaction (reference: Highest SEP sub-group).

**Figure 5.**
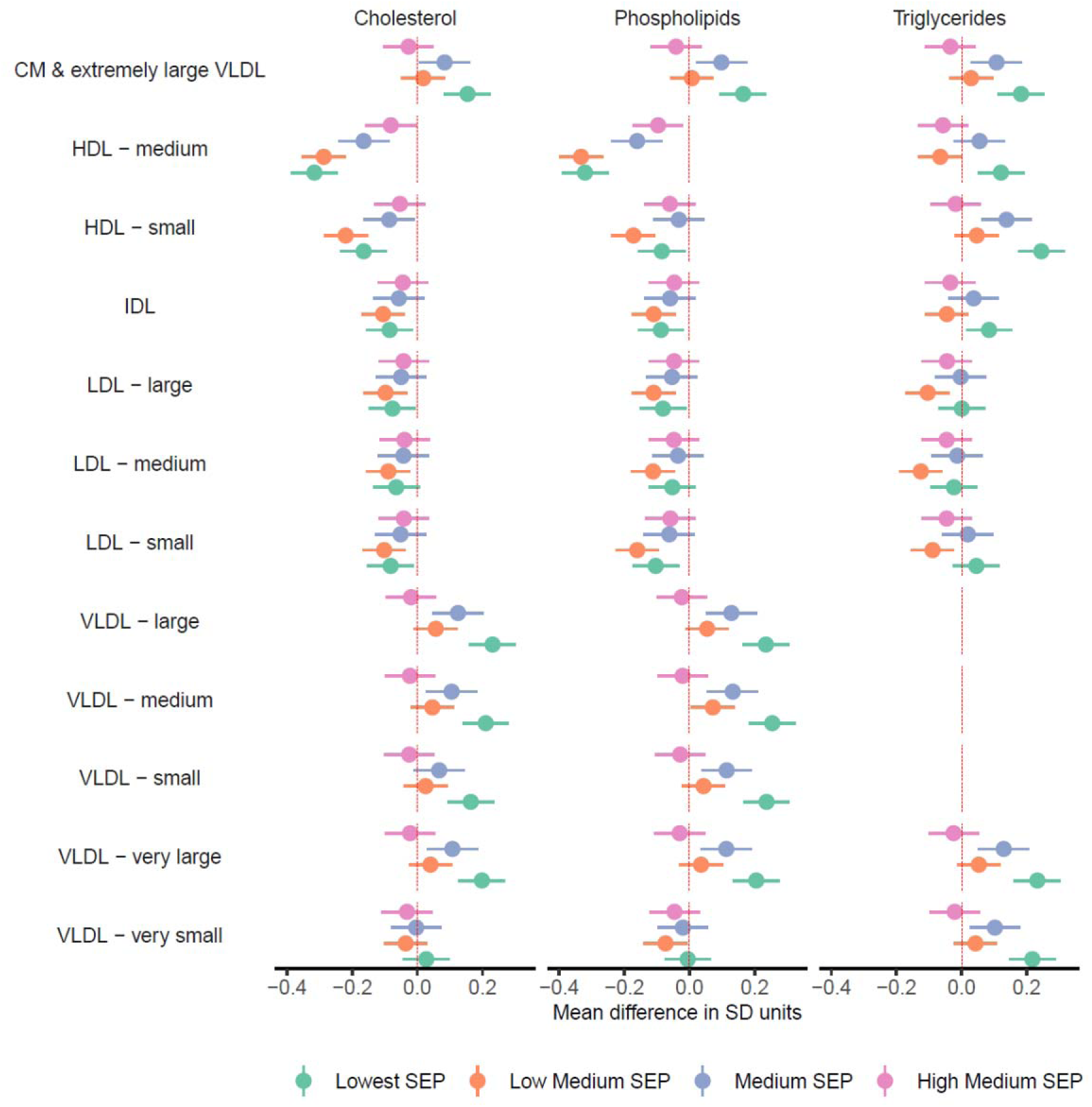
Mean difference in cholesterol, phospholipids, and triglycerides in lipoprotein subclasses by SEP sub-groups in the combined sample of White European and South Asian women, shown for traits without statistical evidence of SEP by ethnicity interaction (reference: Highest SEP sub-group).

There was a statistical interaction between SEP and ethnicity (at the FDR corrected *P* < 0.1 threshold) for 31 metabolic traits (**Additional File 5: Data Set 2**). For most of these traits, differences by SEP group were larger and showed a clearer gradient in White European than South Asian women (**Additional File 7: Data Set 4**). This included stronger association with omega−3 fatty acids, cholesterol and triglycerides in large HDL, docosahexaenoic acid, and degree of unsaturation in White Europeans (**Figure 6**). Differences in HDL and VLDL particle size were larger in White Europeans but the difference in LDL particle size was larger in South Asians (**Additional File 7: Data Set 4**).

**Figure 6.**
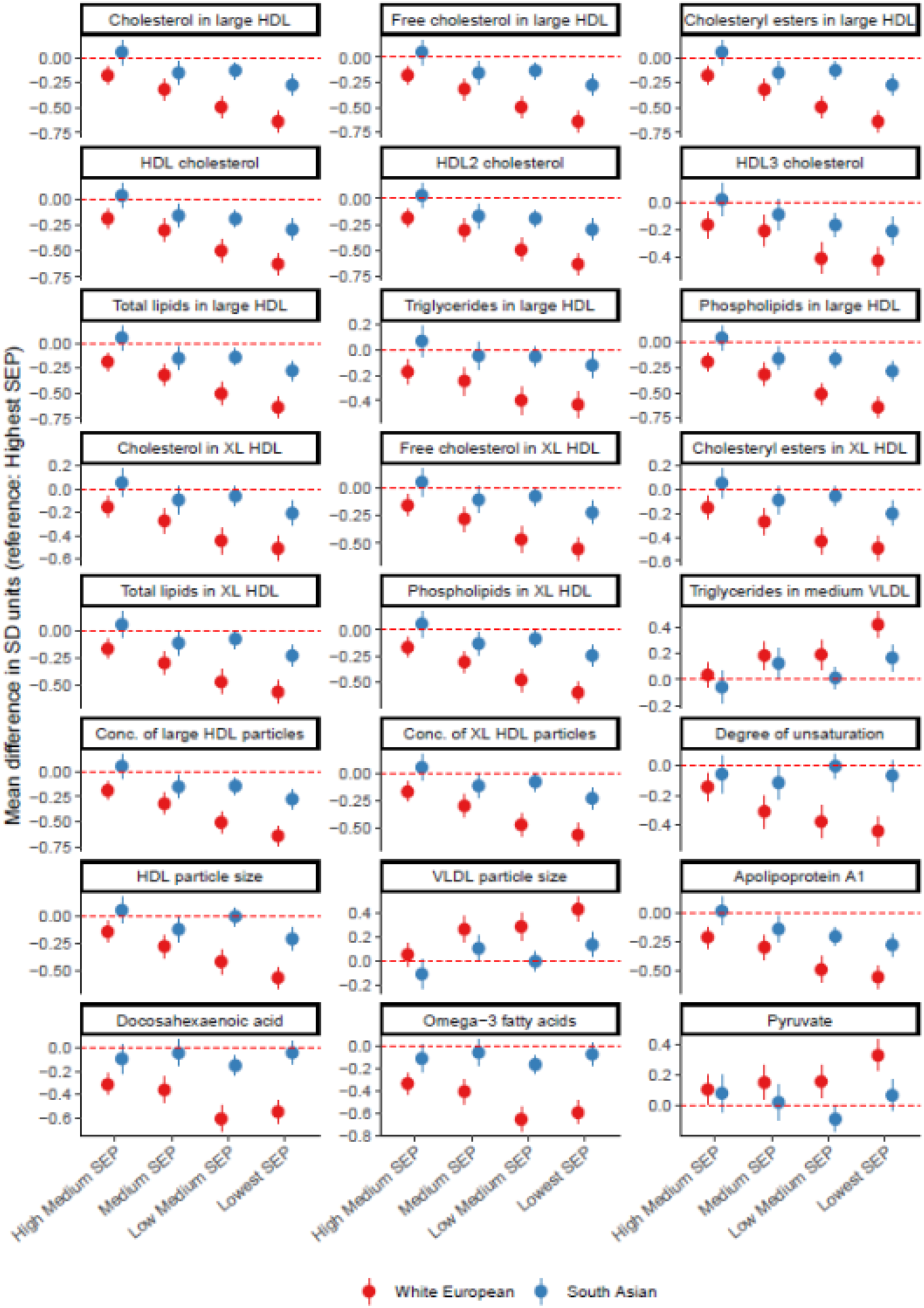
Mean difference in pregnancy metabolic traits by SEP sub-groups in combined sample of White European and South Asian women, presented for top 24 metabolic traits with evidence of SEP by ethnicity interaction (all FDR adjusted P_interaction_ < 0. 05).

### Ethnicity-specific SEP sub-groups and pregnancy metabolic traits

Ethnicity-specific LCA identified five sub-groups in White Europeans and three in South Asians. SEP sub-groups in White Europeans were labelled ‘*Highest SEP’*, ‘*High-Medium SEP’*, ‘*Medium SEP’*, ‘*Low-Medium SEP’*, and ‘*Lowest SEP’*, and SEP sub-groups in South Asians were labelled ‘*Highest SEP’*, ‘*Medium SEP’*, ‘and ‘*Lowest SEP’* (**Figure 7**). As seen for the combined SEP sub-groups, differentiation into sub-groups was driven by a few SEP indicators and there was little difference between sub-groups in whether being able to keep the home warm enough in winter or able to afford two pairs of all-weather shoes (**Figure 7**).

**Figure 7.**
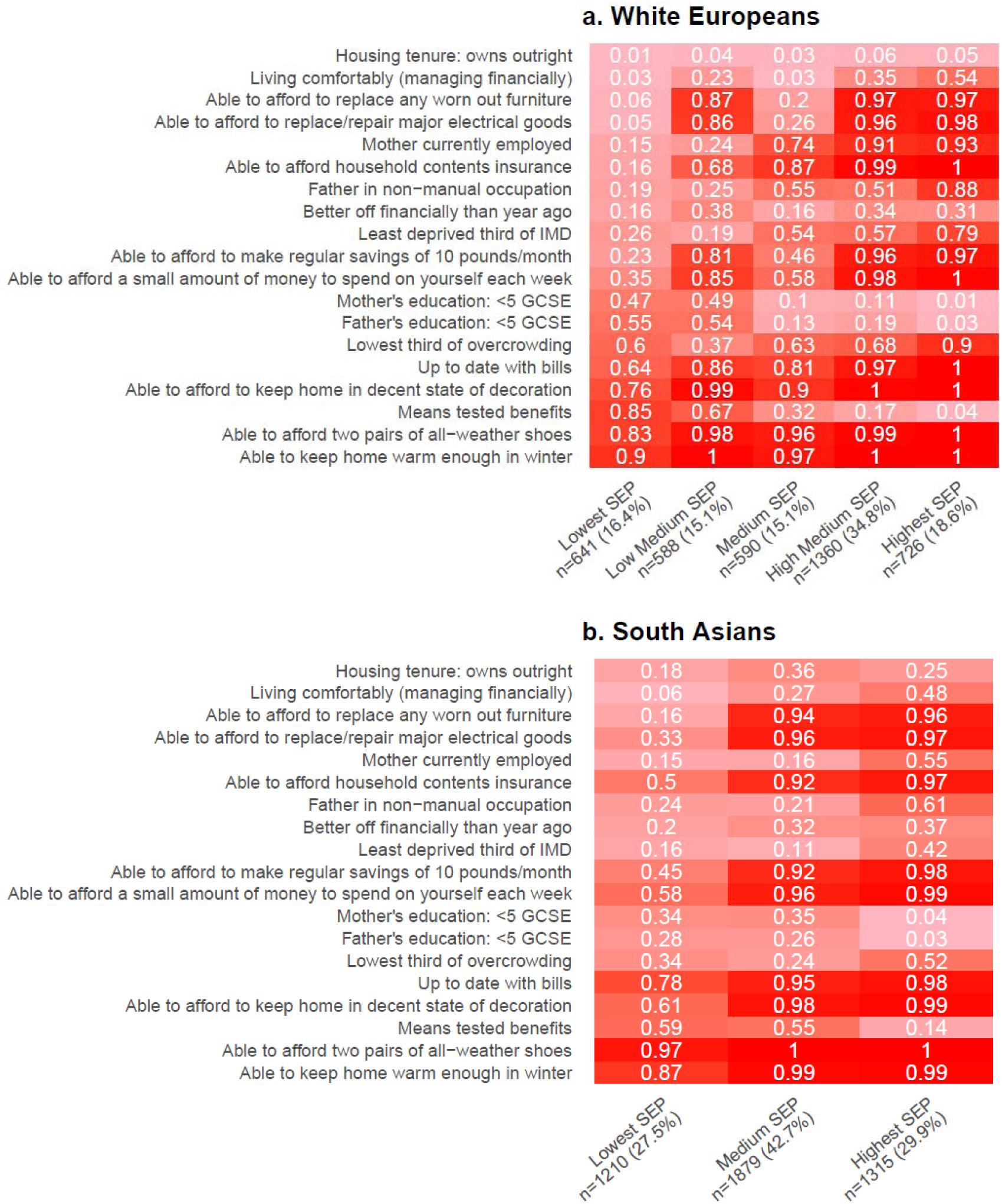
Estimated mean probabilities in each ethnicity-specific SEP sub-group for White European and South Asian women from ethnicity-specific latent class analysis.

Ethnicity-specific SEP was associated with 115 and 98 metabolic traits at the FDR corrected *P*<0.05 threshold in White Europeans and South Asians, respectively (**Additional File 8: Data Set 5**). Results were consistent with those in the combined SEP analysis and included associations in both White Europeans and South Asians between lower SEP and lower HDL-cholesterol and cholines, and higher triglycerides (**Figure 8**). Analyses in White British and Pakistani women identified similar SEP groups, and similar differences in metabolic traits to those found in White Europeans and South Asians, respectively (**Additional File 8: Data Set 5**, **Additional File 9: Figure S2**).

**Figure 8.**
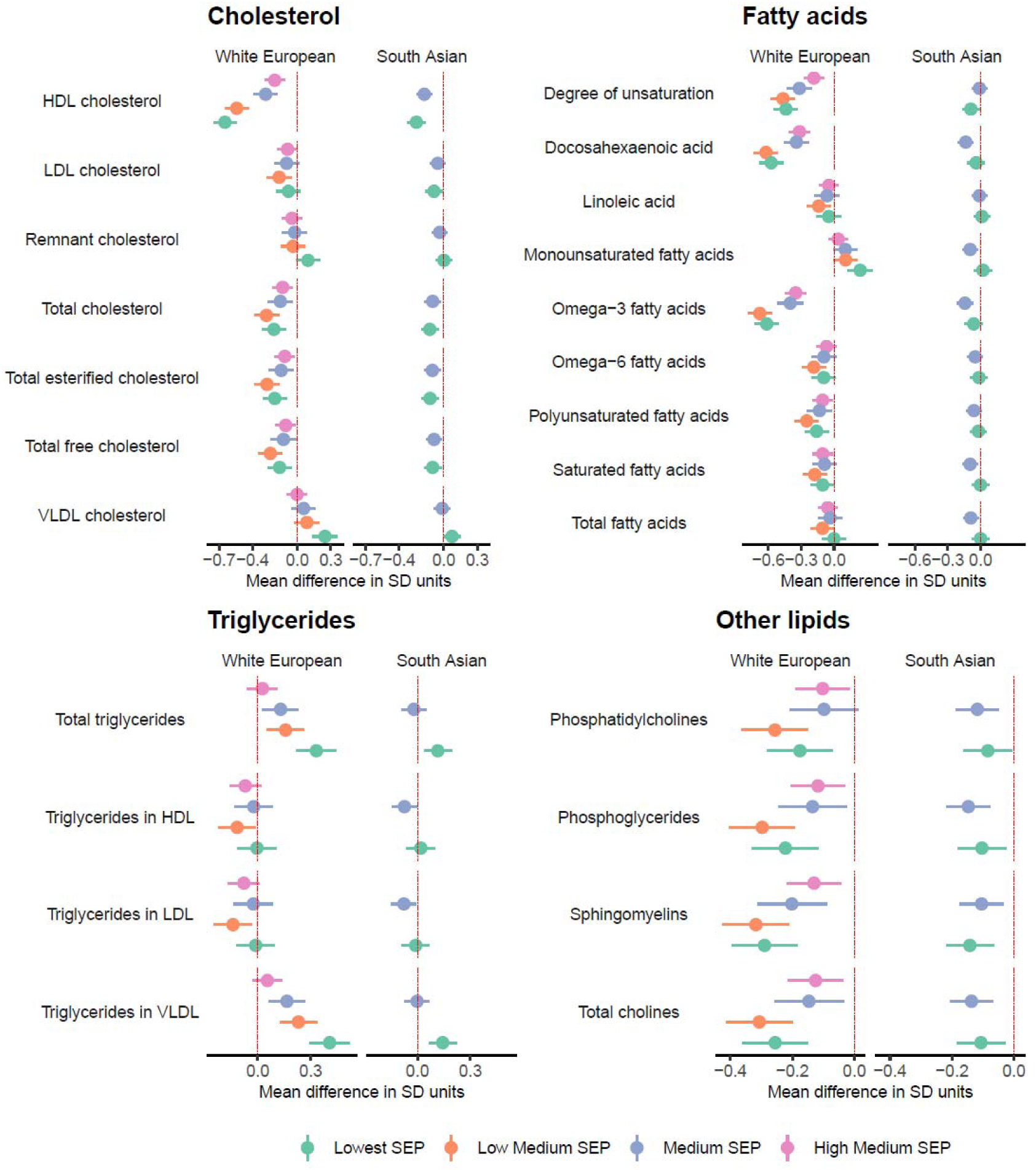
Mean difference in cholesterol, fatty acids, triglycerides, and other lipids by ethnicity-specific SEP sub-groups in White European and South Asian women.

## DISCUSSION

We examined the association between a composite measure of SEP that should better reflect its multidimensional nature and 148 serum metabolic traits in pregnant White European and South Asian women. We found substantial socioeconomic differences across most metabolic traits characterized by more adverse levels of traits in lower SEP subgroups. These included SEP differences across most medium, large, and very large HDL lipoprotein subclasses, and small, medium, large, and very large VLDL subclasses. There was statistical evidence that association with some traits were larger in White Europeans than South Asians, including omega−3 fatty acids, cholesterol and triglycerides in large HDL, docosahexaenoic acid, pyruvate, apolipoprotein A1, degree of unsaturation, and HDL and VLDL particle size.

Our findings are consistent with results from 30 000 adults and 4000 children across 10 UK and Finnish cohort studies which found associations between lower educational attainment and occupational class and more adverse (NMR-derived) metabolic traits including lower HDL traits^10^. We found that lower SEP associated with higher levels of the inflammatory marker glycoprotein acetyls corroborates findings from a study of 605 women showing that higher educational level was associated with lower inflammatory biomarkers in pregnancy^27^. Given that SEP and NMR metabolic traits can inform on adverse pregnancy and perinatal outcomes^4–9^, our findings suggest SEP might influence adverse pregnancy and perinatal outcomes via effects on metabolic traits.

There are complex and multi-factorial factors contributing to socioeconomic differences in health outcomes^28–30^. Individual-level attributes including diet, physical activity, BMI, and smoking are strongly socially patterned and can influence metabolic profiles and therefore are likely to explain some of the socioeconomic differences observed^31–34^. For example, BMI, smoking, HDL-cholesterol and blood pressure have been shown to explain a considerable amount of the association between SEP and adverse pregnancy and perinatal outcomes^35^.

Features of the built environment might also contribute to socioeconomic differences in pregnancy metabolic profiles^36–39^. Socioeconomic differences in metabolic traits might also be attributable to differential developmental trajectories shaped by early life experiences and cumulative allostatic load over the life course^40^

We found statistical evidence to suggest that around 25% of the associations between SEP and metabolic traits were stronger and showed clearer socioeconomic gradient in the White Europeans than South Asians. Given evidence that distributions of most pregnancy NMR metabolic traits differed between White European and South Asians^14^, it is possible that SEP contributes to differences in pregnancy metabolic traits and differences in adverse pregnancy and perinatal outcomes between White Europeans and South Asians. Less variation in risk factors, including health behaviours, between SEP sub-groups among South Asian women might be one explanation for the stronger socioeconomic differences in White Europeans found in our study. For example, South Asian women in the lower SEP groups might have healthier dietary habits, e.g., higher home-prepared food consumption and lower snack consumption^41–43^, and lower smoking rates^44^ than lower SEP White European women. Our ethnicity specific LCA identified fewer SEP sub-groups in South Asians indicating lesser variability in SEP in South Asians, which might also contribute to ethnic differences in associations. One reason for the lower variability in SEP in South Asians might be due to them being mostly first-generation immigrants. New immigrants from the same geographic area tend to be more homogenous in their socioeconomic background and would have not yet established the inequality patterns and socioeconomic gradients of the local population since these require time and acculturation before they emerge in subsequent generations^45^. Finally, difference in perceived adversities and the way to face social adversities could also contribute to explaining these ethnic group difference in how SEP influences metabolic traits^46^

### Limitations

Our study only included White Europeans and South Asians and therefore findings may not generalise to other ethnic groups. The participants were from a high-income country and so findings might not generalise to Whites and South Asians in low-income countries. Women with incomplete data on all SEP indicators were included in LCA using FIML which gives unbiased results under the missing at random assumption. However, if this assumption does not hold, this can produce bias and make the model selection criteria less reliable. We found that only a few SEP indicators explained most of the variation between SEP latent classes therefore, future studies might want to compare LCA to the conventional approach of using one SEP indicator. The Nightingale NMR platform used here primarily covers lipoproteins and therefore we have not assessed other classes of metabolites in detail.

### Conclusions

We found widespread and sizeable socioeconomic differences in metabolic traits in pregnant White European and South Asian women characterized by more adverse levels of metabolic traits in lower SEP subgroups, with statistical evidence of stronger associations for some of the metabolic traits in White European than South Asian women. Our findings suggest that, in the context of limited resources, supporting all pregnant women in lowest SEP groups might provide the greatest benefit for pregnancy and perinatal health.

## Supporting information

Data Set 1

Data Set 3

Data Set 4

Data Set 5

Data Set 2

## Data Availability

The data used during the current study are available to researchers by request from the Born In Bradford Executive Group

## LIST OF ABBREVIATIONS

BiB: Born in Bradford
IMD: Index of Multiple Deprivation
LCA: Latent class analysis
NMR: Nuclear Magnetic Resonance
SEP: Socioeconomic position

## DECLARATIONS

### Ethics approval and consent to participate

BiB had ethical approval from Bradford Research Ethics Committee (07/H1302/112), YFS: Hospital District of Southwest Finland (ETMK:68/1801/2017). All BiB participants provided informed consent or assent to participate in the study and secondary data analyses.

### Consent for publication

Not applicable

### Availability of data and materials

The data used during the current study are available to researchers by request from the BiB Executive Group

### Competing interests

DAL reported grants from national and international government and charity funders, Roche Diagnostics, and Medtronic Ltd for work unrelated to this publication. DAL also declares that she is an editor for BMC Medicine. The other authors report no conflicts.

### Funding

This project has received funding from the European Union’s Horizon 2020 research and innovation programme under grant agreements No. 874583 (ATHLETE), and No. 874739 (LongITools). AE received part of his salary from the European Union’s Horizon 2020 research and innovation programme under grant agreement No. 101021566 (ART-HEALTH). AE, GLC, AGS, KT, and DAL work in a unit that is supported by the University of Bristol and UK Medical Research Council (MC_UU_00011/6). BiB has received funding from the Wellcome Trust (101597), a joint grant from the UK Medical Research Council and UK Economic and Social Science Research Council (MR/N024391/1), and a British Heart Foundation Clinical Study grant (CS/16/4/32482). ISGlobal acknowledges support from the grant CEX2018-000806-S funded by MCIN/AEI/ 10.13039/501100011033, and support from the Generalitat de Catalunya through the CERCA Program. The funders had no role in the design and conduct of the study; management, analysis, and interpretation of data; preparation, review, or approval of the manuscript; and decision to submit the manuscript for publication.

### Authors’ contributions

AE developed the idea for this study with initial input from MV and LM. AE developed the analysis plan with input from all authors. AE undertook all analysis and wrote the first draft of the manuscript. GLC, AGS, KT, LM, GS, NJT, JW, DAL, and MV provided feedback on the draft and approved the final manuscript for submission.

## Acknowledgements

We are grateful to everyone involved in the Born in Bradford study. This includes the families who kindly participated, as well as the practitioners and researchers all of whom made Born in Bradford happen. Sample processing and NMR analysis were carried out at the Bristol Bioresource Laboratory and the NMR Metabolomics facility at University of Bristol.

## SUPPLEMENTAL MATERIAL

**Additional File 1: Data Set 1.** Metabolic traits included in this study.

**Additional File 2: Table S1.** Results of latent class models with 2 to 6 classes.

**Additional File 3: Table S2.** Comparison of study participants with those excluded due to missing data on metabolic traits and gestational age.

**Additional File 4: Figure S1.** Distribution of metabolic traits by ethnicity group

**Additional File 5: Data Set 2.** *P*-values and FDR-adjusted *P*-values for SEP and for SEP by ethnicity interaction term for each metabolic trait.

**Additional File 6: Data Set 3.** Associations between combined SEP latent class sub-groups and standardized pregnancy metabolic trait, presented as mean difference by SEP sub-group (vs. Highest SEP) in the combined sample of White European and South Asian women.

**Additional File 7: Data Set 4.** Associations between combined SEP latent class sub-groups and standardized pregnancy metabolic trait, presented as mean difference by SEP sub-group (vs. Highest SEP) separately in White European and South Asian women.

**Additional File 8: Data Set 5.** Associations between ethnicity-specific SEP latent class sub-groups and standardized pregnancy metabolic trait in White European, White British, South Asian, and Pakistani women.

**Additional File 9: Figure S2**. Estimated mean probabilities for each SEP indicator variable in each ethnicity-specific SEP latent class sub-group in White British and Pakistani women.

## Notes

### Clinical Protocols

https://osf.io/xrf8s/

### Funding Statement

This project has received funding from the European Unions Horizon 2020 research and innovation programme under grant agreements No. 874583 (ATHLETE) and No. 874739 (LongITools). AE received part of his salary from the European Unions Horizon 2020 research and innovation programme under grant agreement No 101021566 (ART-HEALTH). AE, GLC, AGS, KT, and DAL work in a unit that is supported by the University of Bristol and UK Medical Research Council (MC_UU_00011/6). BiB has received funding from the Wellcome Trust (101597) a joint grant from the UK Medical Research Council and UK Economic and Social Science Research Council (MR/N024391/1) and a British Heart Foundation Clinical Study grant (CS/16/4/32482). ISGlobal acknowledges support from the grant CEX2018-000806-S funded by MCIN/AEI/ 10.13039/501100011033, and support from the Generalitat de Catalunya through the CERCA Program. The funders had no role in the design and conduct of the study management analysis and interpretation of data preparation review or approval of the manuscript and decision to submit the manuscript for publication.

### Author Declarations

The Born In Bradford study had ethical approval from Bradford Research Ethics Committee (07/H1302/112), YFS: Hospital District of Southwest Finland (ETMK:68/1801/2017). All Born In Bradford sudy participants provided informed consent or assent to participate in the study and secondary data analyses.

