## Supplementary figures and images for "Social inequalities in pregnancy metabolic profile: findings from the multi-ethnic Born in Bradford cohort study"

### Figure S1

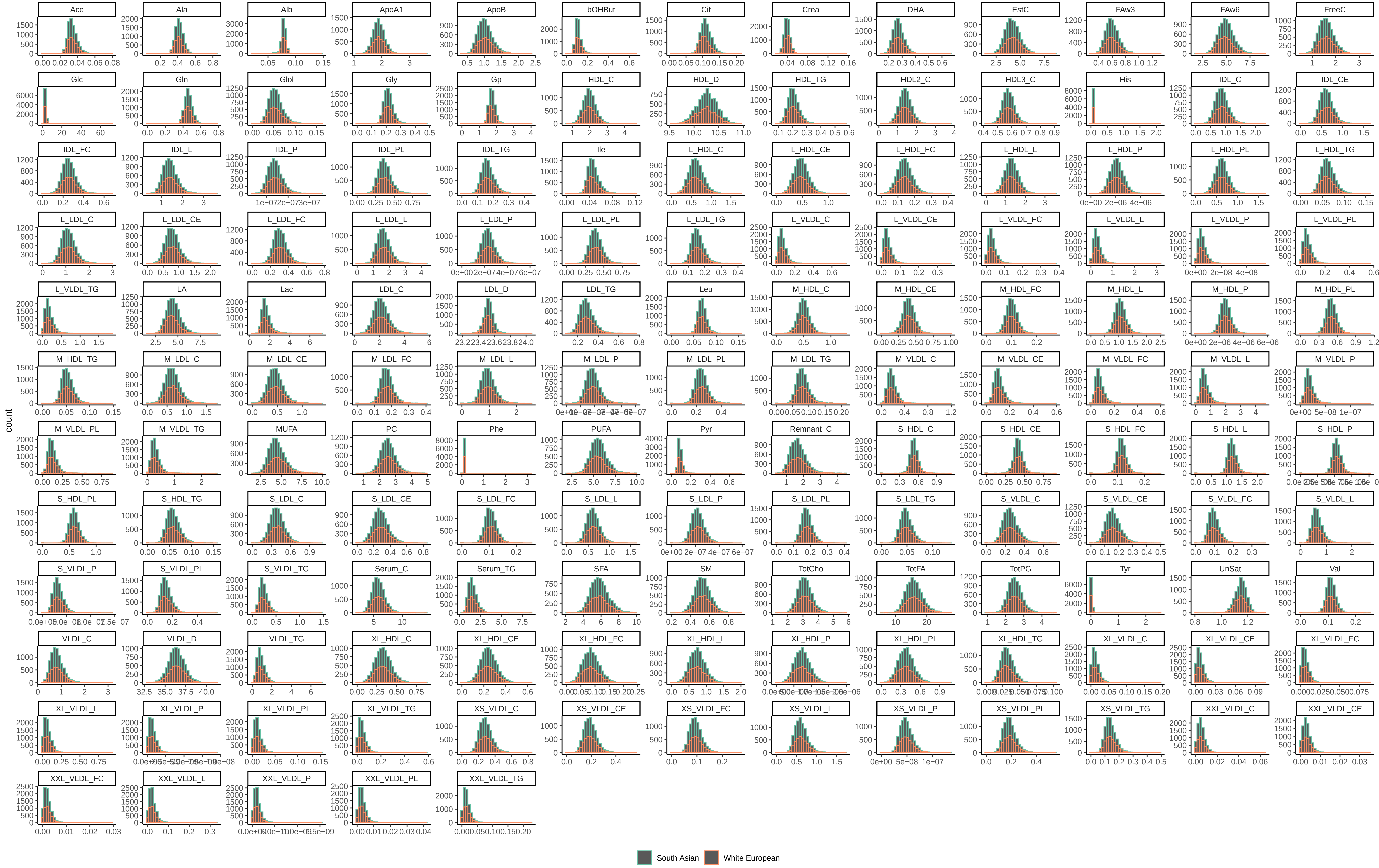

### Figure S2

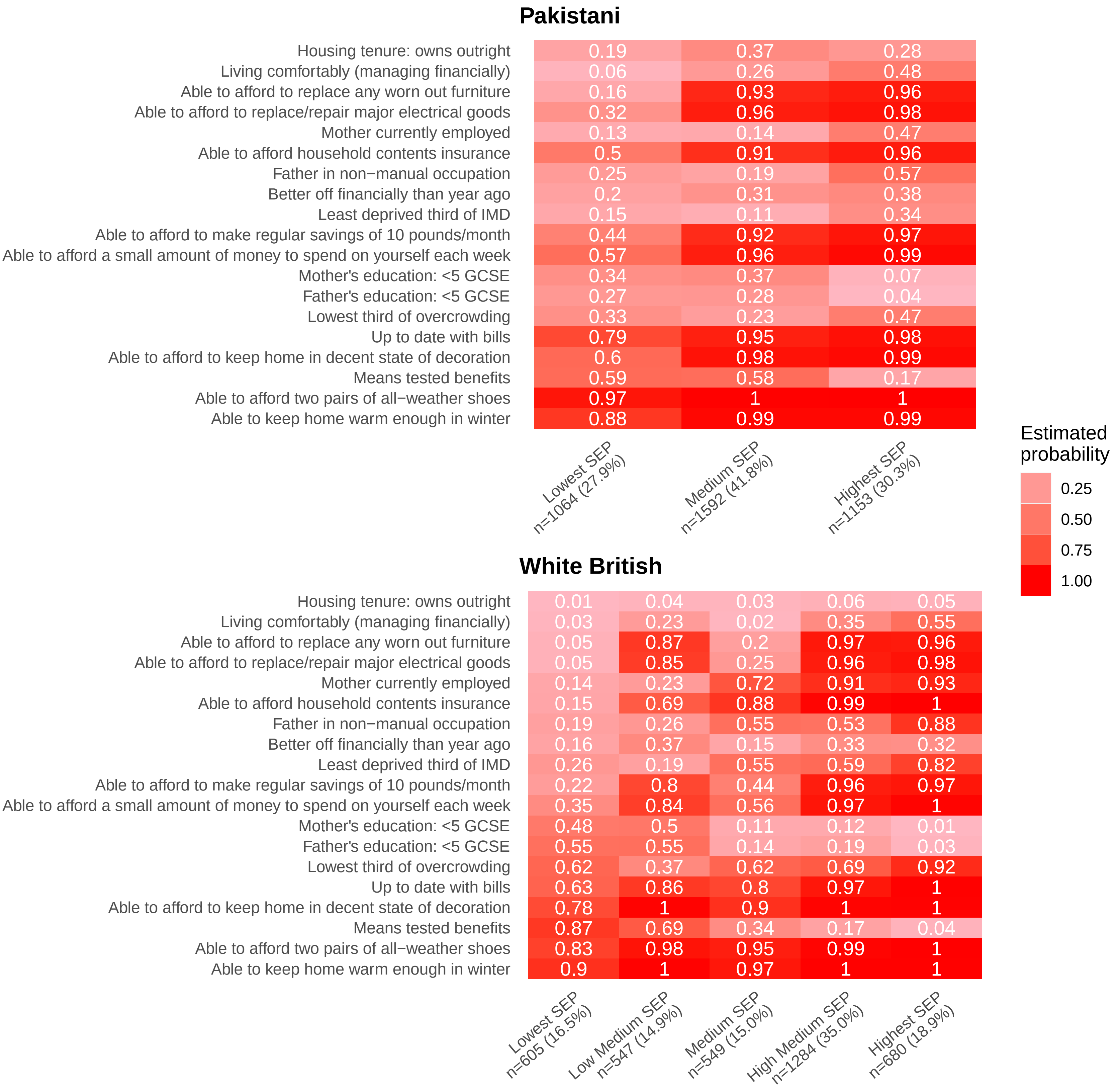
