## Supplementary material for "Social inequalities in pregnancy metabolic profile: findings from the multi-ethnic Born in Bradford cohort study": Table S1

**Additional File 2: Table S1.** Results of latent class models with 2 to 6 classes

| Model | BIC | LMR | Entropy | Class size (%) |
| --- | --- | --- | --- | --- |
| *White European and South Asian* |  |  |  |  |
| k = 2 | 211332 | P<0.0001 | 0.85 | 5388 (64.8), 2921 (35.2) |
| k = 3 | 207011 | P<0.0001 | 0.79 | 3138 (37.8), 2824 (34.0), 2347 (28.2) |
| k = 4 | 205778 | P<0.0001 | 0.78 | 2815 (33.9), 2790 (33.6), 1662 (20.0), 1042 (12.5) |
| k = 5 | 205063 | P<0.0001 | 0.77 | 2623 (31.6), 1627 (19.6), 1526 (18.4), 1460 (17.6), 1073 (12.9) |
| k = 6 | 204584 | P=0.5 | 0.75 | 2182 (26.3), 1617 (19.5), 1491 (17.9), 1459 (17.6), 864 (10.4), 696 (8.4) |
| *White European* |  |  |  |  |
| k = 2 | 92589 | P<0.0001 | 0.881 | 2367 (60.6), 1538 (39.4) |
| k = 3 | 90849 | P<0.0001 | 0.835 | 1955 (50.1), 1088 (27.9), 862 (22.1) |
| k = 4 | 89853 | P<0.0001 | 0.829 | 1776 (45.5), 832 (21.3), 692 (17.7), 605 (15.5) |
| k = 5 | 89443 | P<0.0001 | 0.793 | 1360 (34.8), 726 (18.6), 641 (16.4), 590 (15.1), 588 (15.1) |
| k = 6 | 89328 | P=0.2 | 0.788 | 1295 (33.2), 709 (18.2), 613 (15.7), 548 (14.0), 475 (12.2), 265 (6.8) |
| *South Asian* |  |  |  |  |
| k = 2 | 112418 | P<0.0001 | 0.831 | 2964 (67.3), 1440 (32.7) |
| k = 3 | 110784 | P<0.0001 | 0.757 | 1879 (42.7), 1315 (29.9), 1210 (27.5) |
| k = 4 | 110556 | P=0.6 | 0.727 | 1447 (32.9), 1206 (27.4), 1133 (25.7), 618 (14.0) |
| k = 5 | 110442 | P=0.2 | 0.728 | 1372 (31.2), 1144 (26.0), 924 (21.0), 659 (15.0), 305 (6.9) |
| k = 6 | 110348 | P=0.05 | 0.710 | 1320 (30.0), 971 (22.0), 649 (14.7), 646 (14.7), 519 (11.8), 299 (6.8) |
| *White British* |  |  |  |  |
| k = 2 | 86542 | P<0.0001 | 0.885 | 2172 (59.3), 1493 (40.7) |
| k = 3 | 84859 | P<0.0001 | 0.840 | 1834 (50.0), 1017 (27.7), 814 (22.2) |
| k = 4 | 83920 | P<0.0001 | 0.835 | 1689 (46.1), 765 (20.9), 654 (17.8), 557 (15.2) |
| k = 5 | 83481 | P<0.0001 | 0.800 | 1284 (35.0), 680 (18.9), 605 (16.5), 549 (15.0), 547 (14.9) |
| k = 6 | 83412 | P=0.4 | 0.795 | 1238 (33.8), 657 (17.9), 568 (15.5), 506 (13.8), 453 (12.4), 243 (6.6) |
| *Pakistani* |  |  |  |  |
| k = 2 | 97618 | P<0.0001 | 0.831 | 2561 (67.2), 1248 (32.8) |
| k = 3 | 96530 | P<0.0001 | 0.736 | 1592 (41.8), 1153 (30.3), 1064 (27.9) |
| k = 4 | 96352 | P=0.08 | 0.714 | 1192 (31.3), 1039 (27.3), 973 (25.5), 605 (15.9) |
| k = 5 | 96280 | P=0.5 | 0.701 | 1117 (29.3), 902 (23.7), 875 (23.0), 583 (15.3), 332 (8.7) |
| k = 6 | 96211 | P=0.1 | 0.702 | 1123 (29.5), 766 (20.1), 609 (16.0), 607 (15.9), 412 (10.8), 292 (7.7) |

BIC: Bayesian information criteria; LMR: Lo-Mendell-Rubin adjusted likelihood ratio test
