## Supplementary material for "Social inequalities in pregnancy metabolic profile: findings from the multi-ethnic Born in Bradford cohort study": Table S2

**Additional File 3: Table S2.** Comparison of study participants with those excluded due to missing data on metabolic traits and gestational age.

|  | White European | | South Asian | |
| --- | --- | --- | --- | --- |
| Socioeconomic indicator | Included (n=3905) | Excluded (n=530) | Included (n=4404) | Excluded (n=877) |
| Index of Multiple Deprivation [No. (%)] |  |  |  |  |
| Q1 (least deprived) | 1947 (49.9) | 249 (47.1) | 960 (21.8) | 178 (20.3) |
| Q2 | 1118 (28.6) | 147 (27.8) | 1638 (37.2) | 345 (39.3) |
| Q3 (most deprived) | 839 (21.5) | 133 (25.1) | 1805 (41.0) | 354 (40.4) |
| Woman’s education [No. (%)] |  |  |  |  |
| <5 GCSE equivalent | 733 (21.0) | 112 (23.8) | 1057 (25.3) | 214 (25.9) |
| 5 GCSE equivalent | 1308 (37.5) | 154 (32.7) | 1296 (31.0) | 279 (33.7) |
| A-level equivalent | 675 (19.3) | 79 (16.8) | 568 (13.6) | 105 (12.7) |
| Higher than A-level | 776 (22.2) | 126 (26.8) | 1254 (30.0) | 230 (27.8) |
| Baby’s father’s education [No. (%)] |  |  |  |  |
| <5 GCSE equivalent | 663 (24.1) | 90 (25.1) | 650 (18.9) | 126 (18.3) |
| 5 GCSE equivalent | 1000 (36.4) | 109 (30.4) | 1016 (29.5) | 215 (31.2) |
| A-level equivalent | 470 (17.1) | 72 (20.1) | 418 (12.1) | 73 (10.6) |
| Higher than A-level | 615 (22.4) | 88 (24.5) | 1365 (39.6) | 276 (40.0) |
| Woman’s employment status [No. (%)] |  |  |  |  |
| Currently employed | 2572 (65.9) | 334 (63.0) | 1211 (27.6) | 238 (27.2) |
| Previously employed | 989 (25.3) | 154 (29.1) | 1269 (28.9) | 313 (35.8) |
| Never employed | 342 (8.8) | 42 (7.9) | 1915 (43.6) | 324 (37.0) |
| Baby’s father’s employment status [No. (%)] |  |  |  |  |
| Non-manual | 1824 (50.8) | 246 (50.5) | 1433 (34.5) | 264 (31.5) |
| Manual | 1068 (29.7) | 147 (30.2) | 1622 (39.0) | 354 (42.2) |
| Self-employed | 368 (10.2) | 46 (9.5) | 825 (19.8) | 158 (18.9) |
| Unemployed | 332 (9.2) | 48 (9.9) | 279 (6.7) | 62 (7.4) |
| Means tested benefit [No. (%)] |  |  |  |  |
| Yes | 1383 (35,5) | 175 (33.1) | 1911 (43.5) | 376 (43.0) |
| No | 2508 (64,5) | 354 (66.9) | 2478 (56.5) | 498 (57.0) |
| How well mother and partner managing financially [No. (%)] |  |  |  |  |
| Living comfortably | 1036 (26.7) | 150 (28.5) | 1213 (27.7) | 236 (27.1) |
| Doing alright | 1607 (41.3) | 195 (37.0) | 1847 (42.2) | 361 (41.5) |
| Just about getting by | 973 (25.0) | 143 (27.1) | 981 (22.4) | 201 (23.1) |
| Quite difficult or very difficult | 272 (7.0) | 39 (7.4) | 333 (7.6) | 73 (8.4) |
| Financial circumstance compared to year ago [No. (%)] |  |  |  |  |
| Better off | 1101 (28.3) | 160 (30.3) | 1308 (30.1) | 254 (29.5) |
| Worse off | 1007 (25.9) | 137 (26.0) | 717 (16.5) | 155 (18.0) |
| About the same | 1778 (45.8) | 231 (43.8) | 2315 (53.4) | 453 (52.6) |
| Able to afford two pairs of all-weather shoes [No. (%)] |  |  |  |  |
| Yes | 3611 (96.1) | 496 (96.3) | 4269 (99.2) | 846 (99.5) |
| No | 145 (3.9) | 19 (83.7) | 33 (0.8) | 4 (0.5) |
| Able to afford a small amount of money to spend on yourself each week [No. (%)] |  |  |  |  |
| Yes | 2966 (79.8) | 408 (81.0) | 3614 (86.5) | 710 (85.5) |
| No | 751 (20.2) | 96 (19.1) | 565 (13.5) | 120 (14.5) |
| Able to afford to make regular savings of £10 a month [No. (%)] |  |  |  |  |
| Yes | 2697 (74.3) | 364 (75.6) | 3294 (80.8) | 601 (77.2) |
| No | 932 (25.7) | 124 (25.4) | 783 (19.2) | 178 (22.9) |
| Housing tenure |  |  |  |  |
| Owns outright | 152 (4.2) | 25 (5.0) | 1068 (27.9) | 197 (25.4) |
| Mortgage | 1845 (50.7) | 236 (47.3) | 2020 (52.7) | 427 (55.0) |
| Private landlord | 1050 (28.9) | 145 (29.1) | 484 (12.6) | 108 (13.9) |
| Social housing | 590 (16.2) | 93 (18.6) | 261 (6.8) | 44 (5.7) |
| Overcrowding (person per room) ([No. (%)] |  |  |  |  |
| Q1 | 2548 (65.4) | 328 (62.0) | 1547 (35.2) | 307 (35.1) |
| Q2 | 849 (21.8) | 112 (21.2) | 1393 (31.7) | 294 (33.6) |
| Q3 | 500 (12.8) | 89 (16.8) | 1454 (33.1) | 275 (31.4) |
| Up to date with bills [No. (%)] |  |  |  |  |
| Yes | 3369 (88.1) | 454 (88.2) | 3854 (91.4) | 747 (89.1) |
| No | 454 (11.9) | 61 (11.8) | 363 (8.6) | 91 (10.9) |
| Able to afford to replace or repair major electrical goods [No. (%)] |  |  |  |  |
| Yes | 2485 (70.2) | 339 (72.0) | 3096 (79.2) | 616 (78.8) |
| No | 1057 (29.8) | 132 (28.0) | 813 (20.8) | 166 (21.2) |
| Able to keep home warm enough in winter [No. (%)] |  |  |  |  |
| Yes | 3761 (97.8) | 508 (96.6) | 4175 (96.0) | 822 (95.9) |
| No | 84 (2.2) | 18 (3.4) | 172 (4.0) | 42 (4.9) |
| Able to afford to replace any worn out furniture [No. (%)] |  |  |  |  |
| Yes | 2437 (69.1) | 338 (70.7) | 2830 (72.8) | 560 (72.6) |
| No | 1088 (30.9) | 140 (29.3) | 1059 (27.2) | 211 (27.4) |
| Able to afford household contents insurance [No. (%)] |  |  |  |  |
| Yes | 2407 (84.6) | 307 (84.8) | 2184 (82.2) | 407 (79.3) |
| No | 438 (15.4) | 55 (15.2) | 474 (17.8) | 106 (20.7) |
| Able to afford to keep home in decent state of decoration [No. (%)] |  |  |  |  |
| Yes | 3581 (94.3) | 489 (95.0) | 3743 (88.3) | 740 (87.6) |
| No | 215 (5.7) | 26 (5.1) | 497 (11.7) | 105 (12.4) |
